# Circulating Tumor-Derived Extracellular Vesicles Predict Clinical Outcomes in ^11^C Choline-identified Oligometastatic Castration-Refractory Prostate Cancer Treated with Stereotactic Ablative Radiotherapy

**DOI:** 10.1101/2021.10.23.21265236

**Authors:** F. Lucien, Y. Kim, J Qian, H. Zhang, A Arafa, F. Abraha, I. Thapa, E. J. Tryggestad, W. S. Harmsen, J. Kosti, H. Ali, J. J. Orme, V. J. Lowe, G. B. Johnson, E. D. Kwon, H. Dong, S. S. Park

## Abstract

Stereotactic ablative radiotherapy (SABR) has demonstrated clinical benefit in oligometastatic prostate cancer patients. However, the risk of developing new distant metastatic lesions remains high and only a minority of patients experience durable progression-free response. Therefore, there is a critical need to identify which subset of oligometastatic patients will benefit from SABR alone versus combination SABR and systemic agents. Herein, we provide the first proof-of-concept for the clinical value of circulating prostate cancer-specific extracellular vesicles (PCEVs) as non-invasive predictor of oncological outcomes in oligometastatic castration-refractory prostate cancer (oCRPC) patients treated with SABR.

We have defined PCEVs and analyzed their kinetics in the peripheral blood of 79 oCRPC patients with nanoscale flow cytometry at baseline and days 1, 7, and 14 post-SABR. High PCEV levels at baseline was predictive of shorter time to distant recurrence (3.5 vs 6.6 months, p=0.0087). Following SABR, PCEV levels reached a peak at day 7 and median overall survival was significantly longer in patients with higher PCEV levels (32.7 vs 27.6 months, p=0.003). This suggests that pre-treatment PCEV levels can be a biomarker of tumor burden while early changes post-treatment can predict response to SABR. In contrast, radiomic analyses of ^11^C-choline PET/CT before and after SABR was not predictive of clinical outcomes. Interestingly, a correlation was noted between PCEV levels and peripheral tumor-reactive CD8 T cells (T_TR_; CD8^+^ CD11a^high^).

This original study demonstrates that circulating PCEVs can serve as prognostic and predictive marker to SABR in order to identify “true” oCRPC patients. In addition, we provide novel insights in the global crosstalk mediated by PCEVs between tumors and immune cells which leads to systemic suppression of immunity against CRPC. This work lays the foundation for future studies that investigate the underpinnings of metastatic progression and providing new therapeutic targets (e.g PCEVs) to improve SABR efficacy and clinical outcomes in treatment resistant CRPC.

## Introduction

Patients with metastatic castration-resistant prostate cancer (mCRPC) whose disease is resistant to chemotherapy and next generation anti-androgen therapy have meager median survival of 13.6 months^1^. A subset of patients presenting with oligometastatic disease, usually defined as ≤ 5 lesions, are ideal candidates for stereotactic ablative radiotherapy (SABR)^2^. Therefore, SABR constitutes a stepping-stone towards the treatment of oligometastatic disease ^3–8^. In two recent phase II randomized trials, SABR was proven to prolong progression-free survival of oligometastatic castration-sensitive prostate cancer with minimal toxicities compared to surveillance^4,7,9^. However, distant failure after SABR remains the primary manifestation of disease progression^7,10^. In OPeRATIC (Oligometastatic Prostate cancer Radiotherapy Augmenting T Immune Cells) phase II trial of oligometastatic CRPC (≤ 3 lesions) identified on ^11^C Choline PET/CT, our group has recently shown that SABR is very effective for local control (75% at 2 years) ^11^. In contrast, median time to distant recurrence was 5.1 months and 19% of patients showed distant recurrence within 3 months which highlights that advanced PET imaging may be insufficient in the identification of “true” oligometastatic patients who benefit from metastasis-directed therapy. To overcome this limitation, the combination of PET imaging with minimally-invasive biomarkers has the potential to improve the selection of true oCRPC patients^12^.

The long-term benefit from SABR in a subset of oligometastatic prostate cancer patients can also be attributed to the induction of a systemic antitumor immune response^7,13–16^. Peripheral expansion of clonotypic and tumor-reactive CD8 T cells is a prerequisite for local antitumor response and abscopal effect^7,11,13^. In line with this, there is a critical need to understand the underpinnings of disease progression and systemic suppression of anti-prostate cancer immunity in patients treated with SABR. This will provide a rationale to develop effective combination therapies as seen in melanoma and lung cancer^17,18^.

Extracellular vesicles (EVs) are emerging as promising liquid biomarkers for cancer diagnosis, prognosis and prediction of treatment response^19^. EVs are nanosized vesicles released by all cell types including tumor cells. They contain surface molecules and cargo from donor cells and they can travel in body fluids such as blood and urine. The clinical utility of EVs in the management of prostate cancer has been an active area of investigation, in particular with risk stratification^20–23^. In contrast, clinical data supporting a potential role of EVs to guide patient selection and monitor response to SABR have not been documented. Herein, we defined and examined the baseline and post-SABR levels of prostate cancer-derived extracellular vesicles (PCEVs) in a cohort of oCRPC patients treated with SABR. We evaluated their performance as marker of disease burden and predictor of response to SABR. Finally, we investigated the association of PCEVs and peripheral CD8 T cells to gain novel biological insights on the crosstalk between tumor and immune cells in response to radiotherapy.

## Materials and Methods

### Patient Cohort

Between August 2016 and December 2019, 89 oligometastatic castration-refractory prostate cancer patients identified with ^11^C-Choline PET/CT were treated with SABR in our institution (NCT02816983; OPeRATIC)^11^. Patients with ≤ 3 extracranial lesions, testosterone levels < 50 ng/dL on ADT, ECOG 0-2, and > 6 months of life expectancy were eligible. Study approval was granted by Mayo Clinic Institutional Review Board (IRB #16-000785). The study was conducted in accordance with the Declaration of Helsinki, and written informed consent was obtained from all participants prior to enrollment. Whole blood was collected at baseline and at three timepoints post-SABR (Day 1, Day 7 and Day 14). 79 of 89 patients provided blood at baseline. Out of 79 patients, a total of 66 patients had their blood drawn at baseline and Day 7 post-SABR. A total of 53 patients provided blood for all timepoints. A separate cohort of 40 wide-spread metastastic CRPC (mCRPC) patients with metastatic lesions detected by conventional CT and/or bone scan and with a PSA level above 2.0 ng/ml was used to compare PCEV levels between oCRPC and mCRPC patients (IRB #21-004451).

### Labeling of Prostate Cancer-Derived Extracellular Vesicles (PCEVs)

PCEVs were labeled using the following monoclonal antibodies: Alexa Fluor 647 conjugated PSMA (3E7 clone, Creative Biolabs) and Alexa Fluor 488 conjugated STEAP1 (SMC1 clone, Mayo Clinic Antibody Hybridoma Core) antibodies. PSMA and STEAP1 antibodies were conjugated using protein labeling kits (A10235 and A20173, Thermo Scientific). Degree of antibody labeling (DOL) was measured using a Nanodrop One C spectrophotometer (Fisher Scientific). DOL for PSMA and STEAP1 was 3.2 and 3.6 respectively. Antibody sensitivity and specificity was validated *in vitro* by using the human prostate cancer LNCaP (PSMA^+^STEAP1^+^), PC3-flu (PSMA^-^STEAP1^-^) and PC3-PIP (PSMA^+^STEAP1^-^) cell lines. PC3-flu and PC3-PIP cell lines were kindly provided by Dr. Xinning Wang (Case Comprehensive Cancer Center, USA). PSMA and STEAP1 protein expression in cell lines was validated by western-blot (**Figure S1A and S1B**). Cell lines were cultured in serum-free media for 24 hours and conditioned medium was collected and concentrated using ultrafiltration (Amicon Ultra-15 100 kDa, Millipore). Positive detection of PSMA^+^- and STEAP1^+^-EVs was analyzed by nanoscale flow cytometry (**Figure S1C and S1D**). Optimal antibody concentrations were determined by titration using three plasma samples with detectable levels of PSMA^+^- and STEAP1^+^-EVs.

### Nanoscale Flow Cytometry

Platelet-poor plasma (PPP) samples were thawed at 37^∘^C for 1 minute and centrifuged at 13,000 x g for 5 minutes at room temperature. PPP was diluted in sterile PBS filtered 0.22 µm and incubated for 30 minutes at room temperature with fluorescently labeled PSMA and STEAP1 antibodies. Labeled samples were further diluted in sterile PBS prior to analysis by nanoscale flow cytometry. Each plasma sample was analyzed on Apogee A60-Micro Plus (Apogee FlowSystems Inc., Northwood, UK) equipped with 3 excitation lasers (405, 488, 638 nm) and 9 detectors. Side scatter (LALS) was used as a trigger at 405 nm laser wavelength. Particle detection by A60-Micro Plus was calibrated with Rosetta calibration beads according to manufacturer’s instructions (Exometry Inc). Before each run, a blank sample with DPBS was run to ensure a count rate < 100 events per second. For sample run, event rate was kept below 7,700 events per second to avoid swarm effect^24^. Each sample was run in three technical replicates for 1 minute and coefficient variation was kept below 15%. Data analysis was performed in FlowJo vs 10.6.1. Number of detected events, sample dilution, flow rate and acquisition time were used to determine particle concentration (EVs per milliliter). For a detailed description of the flow cytometer specifications, pre-analytical and analytical procedures, please refer to the MIFlowCyt-EV report (**Supplementary Materials**).

### PET/CT Imaging Data Analysis

^11^C Choline positron emission tomography (PET) and computed tomography (CT) have been performed for all the patients. Quantitative image features extracted from PET/CT images include: region of interest (ROI) Volume, ROIMaxHU, ROIMeanHU, ROIMaxSUV, ROIMeanSUV, TotalVolume and TotalGlycolysis. These seven features were chosen as they are likely to correlate with tumor/metastasis burden, and can be divided into two groups based on the ROI selected by the algorithm described below and the sum of the total treatment volume.

To select the ROI, the baseline PET/CT were first registered rigidly to the planning CT for each treated site and the CTV_high (Clinical Target Volume) volume (defined by treating physician) was copied to PET/CT. For patient with multiple lesions treated, the maximum SUV of each lesion was compared and volume with the highest SUV was selected as the ROI. The volume of the ROI is calculated as ROIVolume.

To normalize the interpatient variation of the PET SUV and CT HU, a slice of descending aorta (DA) was contoured by a nuclear medicine physician and the mean SUV from PET (DAMeanSUV) and its mean HU from CT (DAMeanHU) were calculated. The ROI based values (ROIMaxHU, ROIMeanHU, ROIMaxSUV, ROIMeanSUV) were normalized by being divided by DAMeanHU and DAMeanSUV, respectively.

TotalVolume and TotalGlycolysis were not based on a selected ROI but on the sum of all treated sites. TotalVolume is the sum of all volume treated. TotalGlycolysis is the sum of the product between mean SUV and the treated volume of each site

### Threshold Optimization

The optimized cut value of each quantitative features was obtained by minimizing the p-value of log rank test with the exhaustive search method. To remove the impact of outliers, for each feature, the 80^th^ percentile and 20^th^ percentiles were used as the maximum and minimum of the search range, which was again divided into 50 intervals (steps) linearly. Starting from the minimum, the cut value was incremented step by step. At each step, the dataset was divided by the cut value into two groups, and the log test p-value for the two groups was calculated if the ratio of the group sizes was between 0.33 and 3.0. The constraint on the ratio of the group sizes was to avoid extreme splitting of the dataset. The cut value with the minimum p-value was taken as the optimized cut value of the feature. To check the stability of the cut, the optimized cut value was shifted by 3% (either in positive direction or negative direction) and the p-value was calculated with the shifted cut value. If the p-new value was shifted by 0.04 or more, the optimized p-value was considered unstable and would be re-optimized. The Python Lifelines module^25^ was utilized for the log rank test and Kaplan-Meier test.

### Immunophenotyping of Peripheral CD8 T Cells

Blood PBMC were isolated and subpopulations were identified with the following antibody panel as previously described^11^: CD8-PE-Cy7 (BD Pharmingen, clone RPA-T8, catalog 304006), CD11a-APC (BioLegend, clone HI111, catalog 301212), PD-1 FITC (BioLegend, clone EH12.2H7, catalog 32990), Bim-PE (Cell Signaling Technology, clone CD34C5, catalog 12186S), Granzyme B-PerCP (Novus Biologicals, clone CLB0GB11, catalog NBP1-50071PCP) and CX3CR1-APC/Cy7 (Biolegend, clone 2A9-1, catalog 341616). Flow cytometry was performed on a CytoFLEX LX (Beckman Coulter, Atlanta, Georgia). Data were analyzed with FlowJo 10.6.1 (Tree Star, Palo Alto, California).

### Statistical Analysis

PSA progression, distant failure and overall survival were used as clinical endpoints to determine the association of PCEV levels with oncological outcomes. Kaplan-Meier estimates were used to estimate survival curves for PSA progression, distant failure and overall survival. For each KM plot, p values were derived from the log-rank test for difference between groups. The hazard ratio (HR) of each biomarker was calculated with univariate Cox proportional hazard model^25^ with different survival targets (PSA progression, distant recurrence and overall survival). To minimize the impact of outliers, the biomarker values were capped by two times of its 95 percentile value and standardized with StandardScaler algorithm in Python scikit-learn package. Association of PCEV levels with clinical features was determined using two-sided Mann-Whitney U tests. Linear regression analysis of PCEV levels with imaging features or levels of peripheral CD8 T cells was used to determine Spearman’s r values and associated p values. For correlative studies, PCEV levels were used as continuous variables. For association with clinical features and oncological outcomes, PCEV levels were converted to categorical variables and used to classify patients as high and low levels. Prism v9.0.1 (GraphPad Software), Python SciPy^26^ and Python scikit-learn^27^ packages were used for all statistical analyses.

## Results

### Kinetics of circulating PCEVs in oCRPC patients treated with SABR

Baseline patient, tumor, and SABR characteristics are summarized in Table 1 and clinical outcomes have been reported previously^11^. Median levels of PSMA^+^-EVs rapidly increased at Day 1 post-SABR by 1.6-fold (95% Cl: 1.2-2.0, p=0.004) reaching 2.1-fold (95% Cl: 1.4-3.0, p<0.0001) increase at Day 7 post-SABR (**Figure 1A**). At Day 14 post-SABR, median levels of PSMA^+^-EVs returned close to baseline (1.2-fold increase 95% Cl: 0.9-1.8, p=0.407).

**Figure 1.**
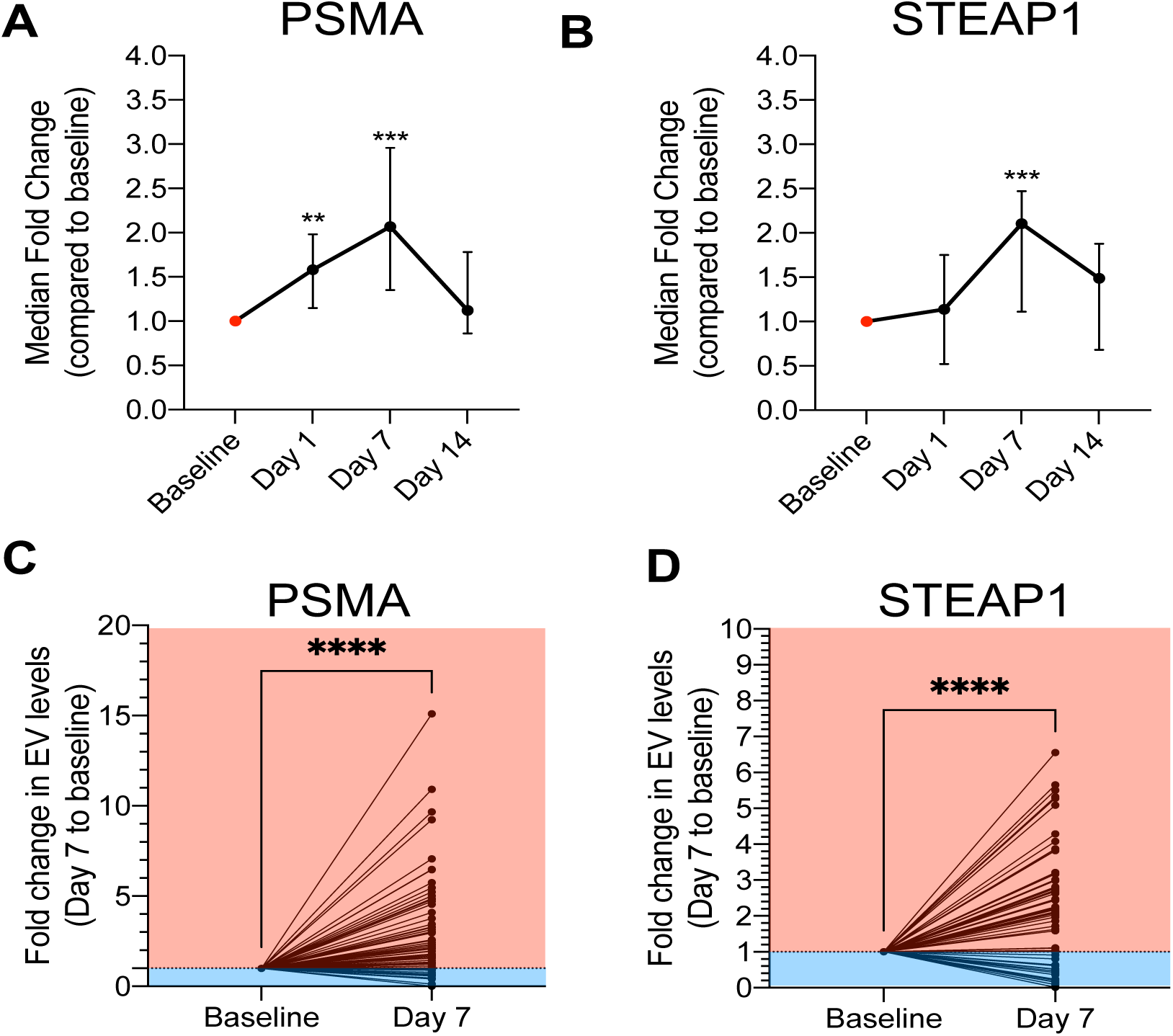
Kinetics of blood levels of PCEVs in response to SABR. A) B) Changes in PSMA^+^-EVs (A) and STEAP1^+^-EVs (B) at 1 day, 7 days and 14 days post-SABR (n=53 pts). Data are presented as median fold change compared to baseline and 95% Cl. Kruskal-Wallis test, **p<0.01, ***p<0.001. C) D) Fold change in levels of PSMA^+^-EVs (C) and STEAP1^+^-EVs (D) between baseline (pre-SABR) and Day 7 post-SABR. Mann-Whitney test, ****p<0.0001, n=66 pts.

**Table 1.**
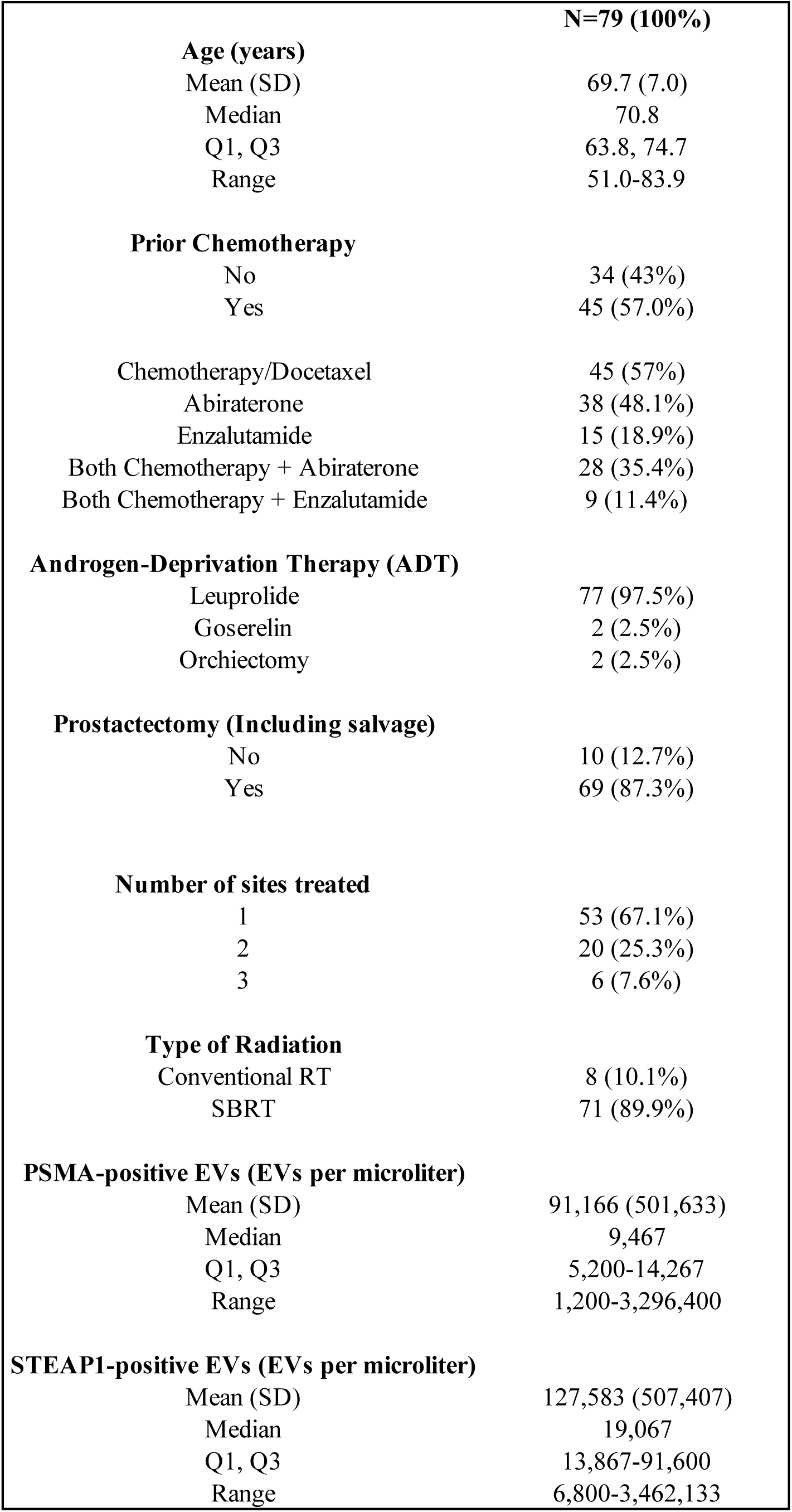
Patient baseline characteristics and demographics.

|  |  |
| --- | --- |
|  | **N=79 (100%)** |
| **Age (years)** |  |
| Mean (SD) | 69.7 (7.0) |
| Median | 70.8 |
| Q1, Q3 | 63.8, 74.7 |
| Range | 51.0-83.9 |
| **Prior Chemotherapy** |  |
| No | 34 (43%) |
| Yes | 45 (57.0%) |
| Chemotherapy/Docetaxel | 45 (57%) |
| Abiraterone | 38 (48.1%) |
| Enzalutamide | 15 (18.9%) |
| Both Chemotherapy + Abiraterone | 28 (35.4%) |
| Both Chemotherapy + Enzalutamide | 9 (11.4%) |
| **Androgen-Deprivation Therapy (ADT)** |  |
| Leuprolide | 77 (97.5%) |
| Goserelin | 2 (2.5%) |
| Orchiectomy | 2 (2.5%) |
| **Prostatectomy (Including salvage)** |  |
| No | 10 (12.7%) |
| Yes | 69 (87.3%) |
| **Number of sites treated** |  |
| 1 | 53 (67.1%) |
| 2 | 20 (25.3%) |
| 3 | 6 (7.6%) |
| **Type of Radiation** |  |
| Conventional RT | 8 (10.1%) |
| SBRT | 71 (89.9%) |
| **PSMA-positive EVs (EVs per microliter)** |  |
| Mean (SD) | 91,166 (501,633) |
| Median | 9,467 |
| Q1, Q3 | 5,200-14,267 |
| Range | 1,200-3,296,400 |
| **STEAP1-positive EVs (EVs per microliter)** |  |
| Mean (SD) | 127,583 (507,407) |
| Median | 19,067 |
| Q1, Q3 | 13,867-91,600 |
| Range | 6,800-3,462,133 |

For STEAP1^+^-EVs, similar trend was observed with a median fold change compared to baseline of 1.1-fold (95% Cl: 0.5-1.8, p=0.180), 2.1-fold (95% Cl: 1.1-2.5, p=0.0009) and 1.5-fold (95% Cl: 0.7-1.9, p=0.168) at Day 1, 7 and 14 post-SABR, respectively (**Figure 1B**). Compared to baseline, levels of PSMA^+^-EVs increased in 51/66 patients (77.2%) and decreased in 15/66 patients (22.8%) at Day 7 post-SABR (**Figure 1C**). Levels of STEAP1^+^-EVs increased in 44/66 patients (66.6%) and decreased in 22/66 patients (33.3%) (**Figure 1D**). Changes in STEAP1^+^-EV levels at Day 7 post-SABR were concordant with changes in PSMA^+^-EVs for 43/66 patients (65.1%).

### Baseline prostate cancer-derived extracellular vesicles (PCEVs) and association with PSA

PSMA^+^ and STEAP1^+^ PCEVs were detectable in all patients, and 74 out of 79 patients (93.7%) had higher circulating STEAP1^+^-EVs than PSMA^+^-EVs at baseline (**Figure 2A**). Median levels of STEAP1^+^-EVs and PSMA^+^-EVs were 19,067 (range: 6,800-3,468,933) and 9,467 (range: 6,800-3,468,933) per microliter of plasma, respectively (**Table 1**). Correlation analysis showed a significant positive correlation between baseline PSMA^+^- and STEAP1^+^-EVs (Spearman r=0.599, 95% Cl: 0.430-0.727, p=5.54 x10^−9^). Serum PSA levels at baseline were significantly higher in patients with high levels of both PSMA^+^-EVs (2.4 vs 0.52 ng/ml, p=0.0018) and STEAP1^+^-EVs (2.7 vs 0.62 ng/ml, p=0.0082) (**Figure S2A**). Correlation analysis using continuous variables showed significant but moderate positive correlation between baseline levels of PSMA^+^-EVs and PSA (Spearman r=0.327, 95% Cl: 0.108-0.516, p=0.003). No significant correlation was observed between serum PSA levels and baseline concentrations of STEAP1^+^-EVs (Spearman r=0.210, 95% Cl: −0.018-0.418, p=0.0628).

**Figure 2.**
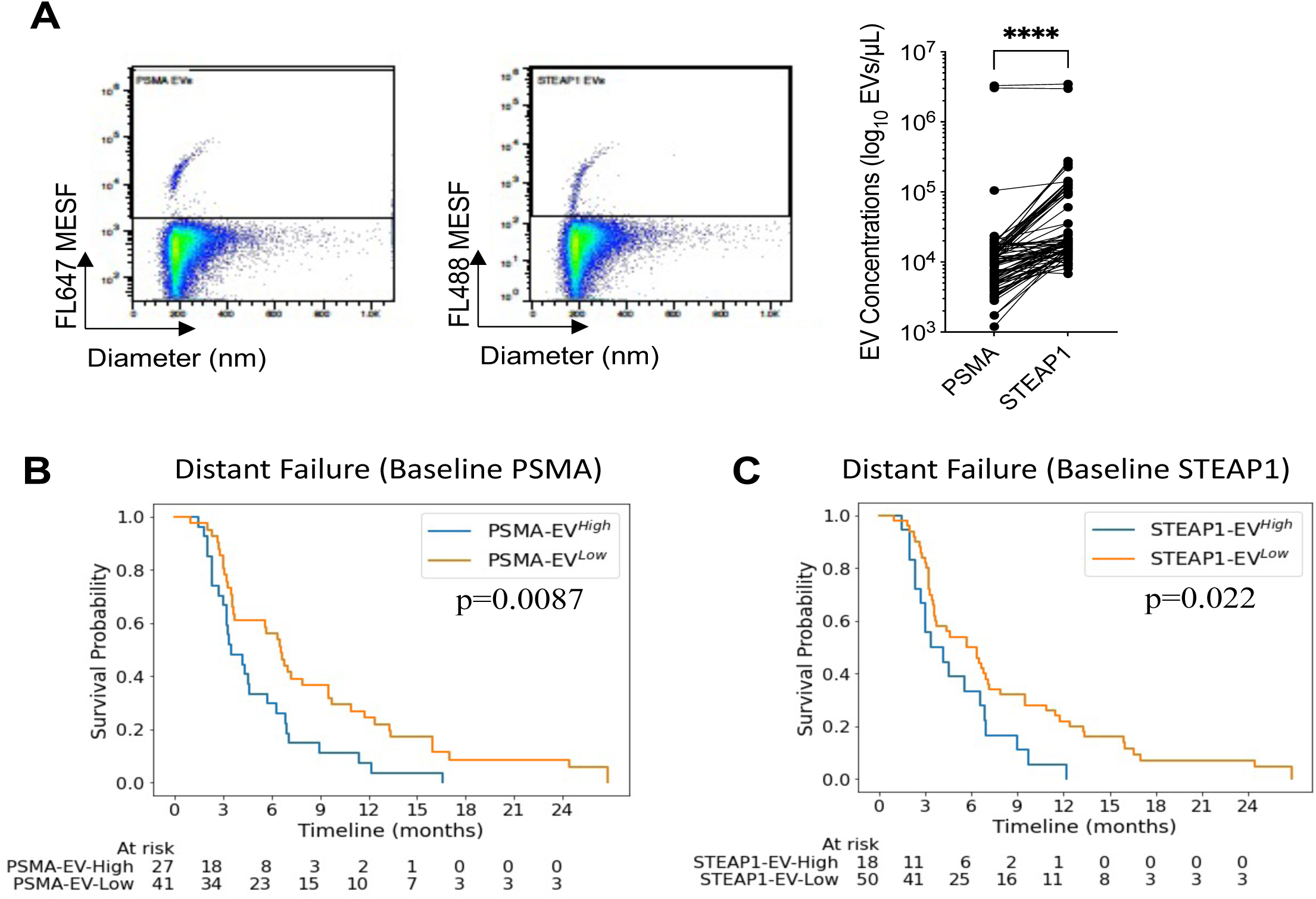
Baseline PCEV levels are risk predictors of distant failure in omCRPC patients treated with SABR. A) Representative scatterplots showing nanoscale flow cytometric detection of PSMA- and STEAP1-positive EVs in patient plasma. Data are presented in logarithmic scale. The dot plot shows baseline concentrations of PSMA- and STEAP1-positive EVs in omCRPC patients (n=79 patients). Wilcoxon matched-pairs signed rank test, ***p<0.000 B) C) Kaplan-Meier estimates of distant failure comparing omCRPC patients who had high and low baseline levels of PSMA^+^-EVs (B) and STEAP1^+^-EVs (C) and treated with SABR. For PSMA^+^-EVs (B), HR=1.35, 95%Cl: 1.03-1.76, p=0.03, n=68. Median time to distant failure for high levels and low levels of PSMA^+^-EVs was 3.47 months and 6.6 months respectively (p=0.0087). For STEAP1^+^-EVs (C), HR=1.43, 95%Cl: 1.09.1.86, p=0.01, n=68. Median time to distant failure for high levels and low levels of STEAP1^+^-EVs was 4.2 months and 5.73 months respectively (p=0.022).

### Baseline PCEVs predict risk of distant failure

Using Cox regression analysis, we analyzed the association of PCEV levels at baseline with PSA progression, distant recurrence, and overall survival (**Table 2**). No association was observed between baseline PCEV levels and PSA progression or overall survival. Both higher baseline PSMA^+^-EVs (HR= 1.35; 95% Cl: 1.03-1.76; p=0.03) and STEAP1^+^-EVs (HR= 1.43; 95% Cl: 1.09-1.86; p=0.01) predict higher risk of distant recurrence independently. Furthermore, high baseline PSMA^+^-EVs or STEAP1^+^-EVs were associated to shorter time to distant recurrence. Median time to distant recurrence was 6.6 months and 3.5 months for patients with low and high levels of PSMA^+^-EVs respectively (p=0.0087) (**Figure 2B**). At 6 months follow-up, distant failure occurred in 19.5% of patients with low PSMA^+^-EVs and 70.4% of patients with high PSMA^+^-EVs. Similarly, median time to distant recurrence was 5.7 months and 4.2 months for patients with low and high STEAP1^+^-EVs respectively (p=0.022) (**Figure 2C**). The risk of distant failure at 6 months post-SABR was 66.6% of patients with high STEAP1^+^-EVs compared to 50% of patients with low STEAP1^+^-EV.

**Table 2.** Association of PCEV levels with oncological outcomes.

| Biomarker | PSA Progression |  |  | Distant Recurrence |  |  | Overall Survival |  |  |
| --- | --- | --- | --- | --- | --- | --- | --- | --- | --- |
|  | n | HR (95% CI) | P value | n | HR (95% CI) | P value | n | HR (95% CI) | P value |
| Baseline PSMA | 79 | 1.21 (0.90 - 1.63) | 0.20 | 63 | **1.35 (1.03 - 1.76)** | **0.03** | 79 | 1.02 (0.70 - 1.47) | 0.93 |
| Baseline STEAP1 | 79 | 1.24 (0.91 - 1.70) | 0.17 | 63 | **1.43 (1.09 - 1.86)** | **0.01** | 79 | 0.91 (0.61 - 1.36) | 0.65 |
| Day 7 PSMA | 75 | 0.71 (0.50 - 1.01) | 0.06 | 57 | **0.68 (0.48 - 0.97)** | **0.03** | 75 | **0.24 (0.07 - 0.80)** | **0.02** |
| Day 7 STEAP1 | 75 | 1.27 (0.99 - 1.62) | 0.06 | 57 | 1.07 (0.82 - 1.40) | 0.64 | 75 | 0.68 (0.40 - 1.18) | 0.17 |
| Change Baseline to Day 7 PSMA | 66 | 0.79 (0.57 - 1.08) | 0.14 | 52 | **0.68 (0.49 - 0.93)** | **0.02** | 66 | 0.54 (0.24 - 1.20) | 0.13 |
| Change Baseline to Day 7 STEAP1 | 66 | 1.17 (0.85 - 1.60) | 0.34 | 52 | 0.81 (0.58 - 1.12) | 0.19 | 66 | 0.89 (0.53 - 1.49) | 0.66 |

### Post-SABR PSMA+-EVs also predict oncological outcomes

Levels of Post-SABR PSMA^+^-EVs at Day 7 were inversely associated with risk of distant recurrence (HR= 0.68; 95% Cl: 0.48-0.97; p=0.03) and overall survival (HR= 0.24; 95% Cl: 0.1-0.8; p=0.02) (**Table 2**). Post-SABR PSMA^+^-EVs were also associated with higher risk of PSA progression (HR= 0.71; 95% Cl: 0.5-1.0; p=0.06). Median time to distant recurrence was 7.0 months and 3.3 months for high and low levels of PSMA^+^-EVs (p=2.9×10^−4^) (**Figure 3A**). At 12 months follow-up, distant failure occurred in 66.6% of patients with high levels of PSMA^+^-EVs and 94.7% of patients with low levels of PSMA^+^-EVs. High levels of Post-SABR PSMA^+^-EVs at day 7 were also associated with longer time to PSA progression (12.8 months vs 6.8 months, p=0.0018) and longer overall survival (32.6 months vs 27.6 months, p=0.003) (**Figure 3B-3C**). Changes in levels of PSMA^+^-EVs from baseline to Day 7 were also predictor of distant recurrence (HR= 0.68; 95% Cl: 0.49-0.93; p=0.02) (**Table 2**). Patients with distant recurrence within 6 months post-SABR had significantly lower median change in PSMA^+^-EV levels compared to patients who did not show distant failure after 12 months (1.1 versus 2.12, p=0.0057) (**Figure 3D**). For STEAP1-positive EVs, no significant association was observed with oncological outcomes.

**Figure 3.**
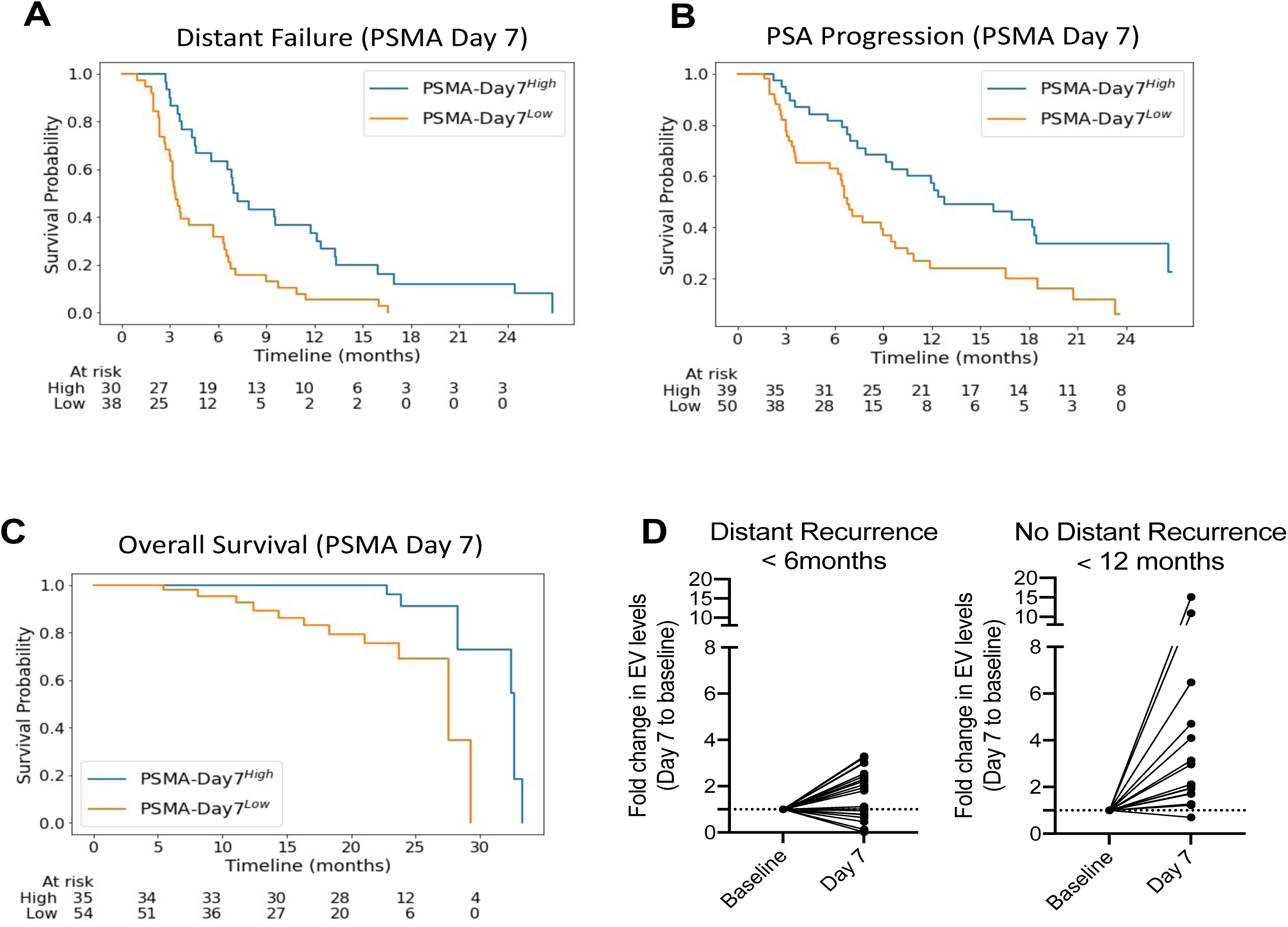
PSMA^+^-EVs levels post-SABR are predictors of oncological outcomes. Kaplan-Meier estimates of distant failure (A), PSA progression (B) and overall survival (C) comparing patients who had high and low baseline levels of PSMA^+^-EVs. C) Fold change in blood levels of PSA^+^-EVs at Day 7 post-SABR in patients presenting distant recurrence within 6 months (left) and no distant recurrence within 12 months (right)

### Association of PCEV levels and immunological changes

Pre-existing antitumor immunity and expansion of tumor-reactive T cells are critical for response to SABR^7,11,28^. In our original study, patients with high levels of tumor-reactive T cells (CD11a^high^ CD8^+^) responded better to SABR with prolonged PSA progression-free survival and time to distant recurrence^11^. An increase in tumor-reactive T cells at day 14 post-SABR was associated with improved overall survival (33.3 months vs 28.2 months; HR 6.21, p=0.04). Previous reports have demonstrated that tumor-derived EVs can carry immunosuppressive proteins and prevent immune-mediated tumor cell killing and response to immunotherapy^29–32^. In line with this, we performed a correlation analysis and evaluate the association between PCEV levels and peripheral CD8 T cells at baseline and post-SABR (**Figure 4A**). At baseline, we found that both levels of PSMA^+^- and STEAP1^+^-EVs were negatively correlated with several subpopulations of tumor-reactive T cells (**Table 3**). No association was found between levels of parent tumor-reactive CD8 T cells (CD11a^high^ CD8^+^) and PCEVs (**Figure 4B**). However, high baseline levels of PSMA^+^- and STEAP1^+^-EVs were associated with lower percentage of tumor-reactive CD8 T cells positive for markers of effector function Bim, CX3CR1/GZMB and PD-1 (**Figure 4C-4D-4E**)^33,34^. Conversely, elevation of PCEV levels at Day 7 was positively correlated with high levels of tumor-reactive CD8 T cells at day 14 post-SABR (**Table 3**) (**Figure 4F**).

**Figure 4.**
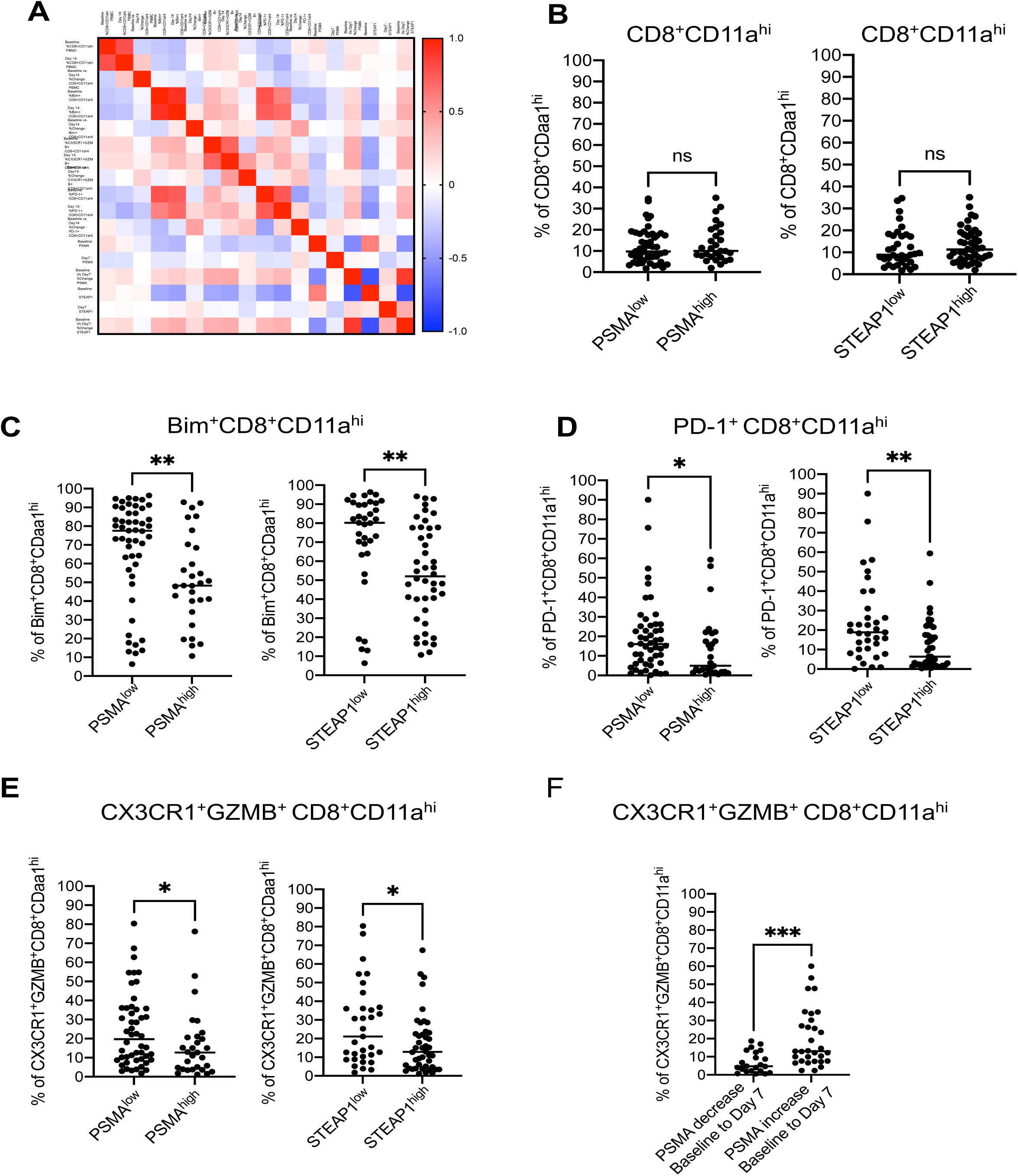
Association of baseline and post-SABR PCEV concentrations with levels of tumor-reactive effector CD8 T cells. A) Correlation matrix of PCEV levels and various subpopulations of peripheral CD8 T cells at baseline and post-SABR. Spearman correlation coefficient is shown from −1.0 (blue) to 1.0 (red). B) C) D) E) Baseline levels of subpopulations of tumor-reactive CD8 T cells in patients stratified by baseline concentrations of PCEVs. Levels of parent tumor-reactive CD8 T cells (B) and populations positive for Bim^+^ (C), PD-1^+^ (D) and CX3CR1^+^GRZB^+^ (E) were compared in patients with PCEV low and high. F) Levels of CX3CR1^+^GRZB^+^ tumor-reactive CD8 T cells at Day 14 in patients with increase or decrease of PSMA^+^-EV concentrations at Day 7 (compared to baseline). Mann-Whitney test, *p<0.05; **p<0.01, ***p<0.001, ns= not significant.

**Table 3.**
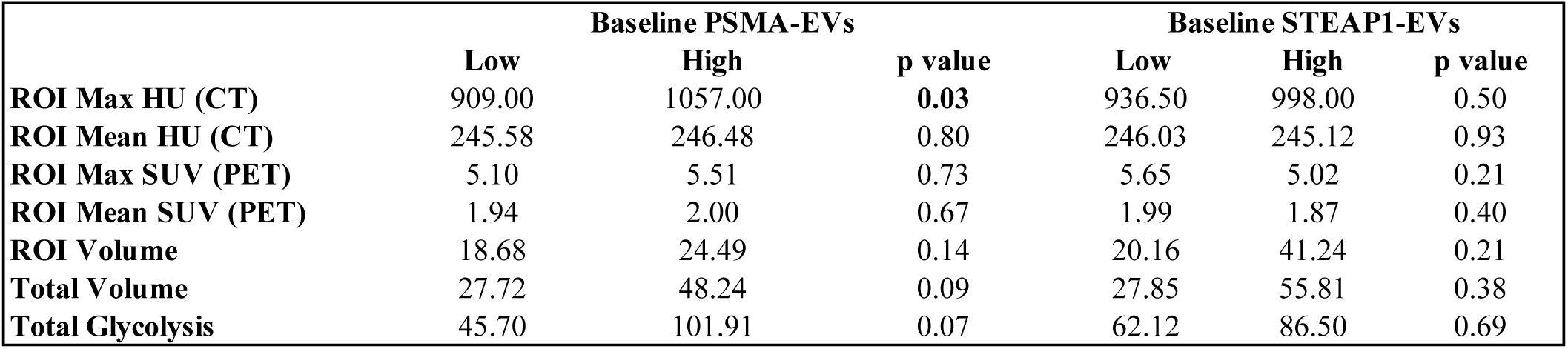
Association of baseline PCEV levels and PET/CT imaging features.

### Association of PCEV levels with tumor burden assessed by ^11^C-Choline PET/CT imaging

In this study, 53/79 (67.1%) had one extracranial metastatic lesion, 20/79 (25.3%) had 2 lesions, and 6/79 (7.6%) had 3 lesions detected by ^11^C-Choline PET/CT imaging (**Table 1**). We analyzed the relationship between baseline PCEV levels and several PET/CT imaging parameters (**Table 4**). No significant association was observed between levels of STEAP1^+^-EVs and tumor volume or characteristics. Higher levels of PSMA^+^-EVs were associated with increased ROI Max HU (CT) (p=0.03), total volume (p=0.09) and total glycolysis (p=0.07). No correlation was found between PCEV levels and imaging features (**Table S1**). No association between PCEV levels and number of metastatic lesions detected on ^11^C-Choline PET/CT imaging was observed (**Figure 5A**). Median levels of PCEVs were similar in patients with solitary metastasis (PSMA: 9,600 EVs per microliter; STEAP1: 20,533 EVs per microliter) than in patients with 2 (PSMA: 8,800 EVs per microliter, STEAP1: 17,867 EVs per microliter) or 3 lesions (PSMA: 8,467 EVs per microliter, 14,267 EVs per microliter). Quantification of STEAP1^+^-EVs shows a wide range of concentrations in patients with 1 or 2 metastatic lesions starting from 8,933 EVs per microliter to 2,973,334 EVs per microliter. We also compared the levels of PSMA^+^-EVs and STEAP1^+^-EVs in oCRPC and mCRPC patients (**Figure 5B**). The cohort of mCRPC is composed of heavily treated patients with widespread metastatic lesions. This cohort presented higher levels of PSMA^+^-EVs compared to oCRPC cohort [median 24,625 EVs per microliter (95% Cl: 17,000-31,750) vs 9,467 EVs per microliter (95% Cl: 7,067-11,600)]. Higher levels of STEAP1^+^-EVs were observed in mCRPC patients with a median level of 105,350 EVs per microliter (95% Cl: 78,500-164,000) vs 19,067 EVs per microliter (95% Cl: 15,467-20,800) for oCRPC patients. Twenty-four of 69 oCRPC patients (30.4%) had STEAP1^+^-EV concentrations in a range of mCRPC patients.

**Figure 5.**
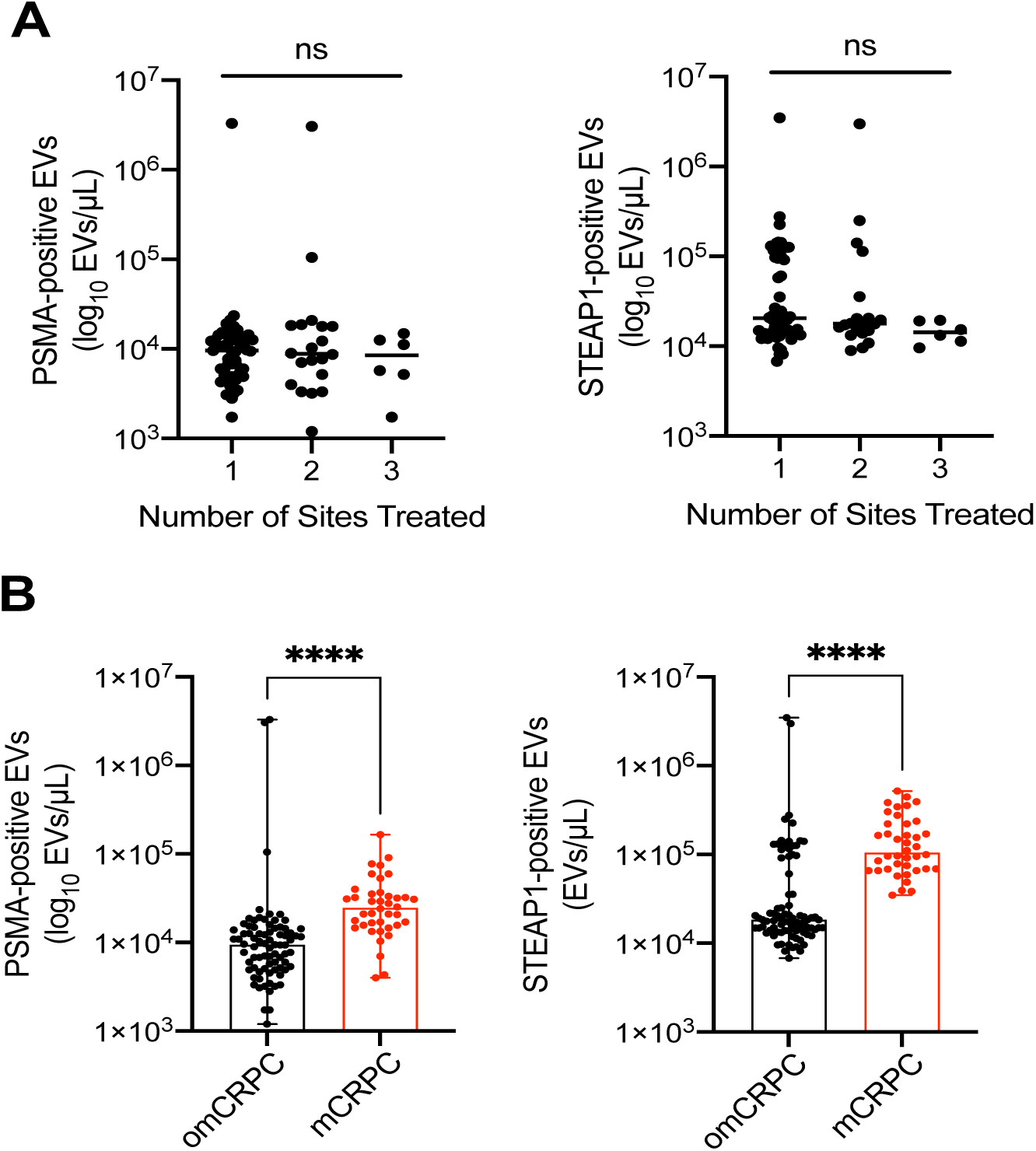
Association of baseline and post-SABR PCEV concentrations with levels of tumor-reactive effector CD8 T cell. A) Baseline levels of PSMA- and STEAP1-positive EVs in patients stratified by number of metastatic lesions identified by ^11^C-Choline PET imaging; 1 lesion (n=53 pts), 2 lesions (n=20 pts), 3 lesions (n=-6). Kruskal-Wallis test, ns= not significant B) Comparison in levels of PSMA- and STEAP1-positive EVs in omCRPC (n=79 pts) and mCRPC patients (n=40 pts). Mann-Whitney test, ***p<0.001.

**Table 4.**
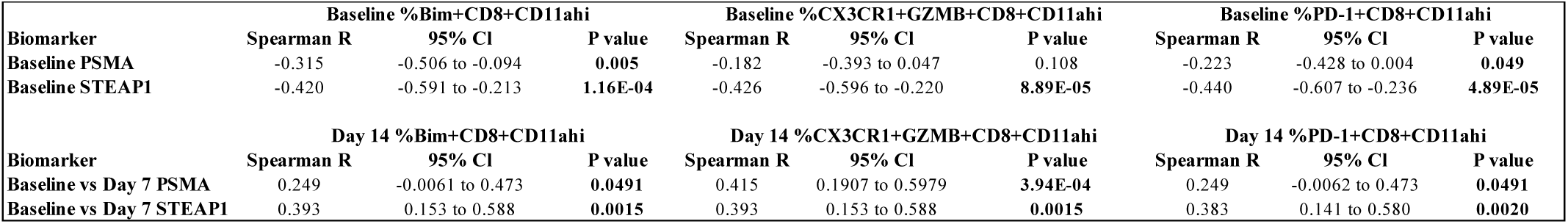
Correlation analysis between PCEV levels and tumor-reactive CD8-T cells.

## Discussion

To our knowledge, this study is the first to analyze the kinetics and evaluate the clinical utility of tumor-derived EVs in prostate cancer patients treated with radiotherapy. It is also the first study to evaluate an EV-based blood test as complementary approach to ^11^C-Choline PET/CT imaging for the diagnosis of oligometastatic prostate cancer. Finally, we analyzed the relationship between circulating EVs and peripheral CD8 T cells at baseline and post-SABR in the intent to provide novel insights in the crosstalk between tumors and immune cells. SABR has shown clinical benefit in oligometastatic prostate cancer patients but defining oligometastatic disease is very challenging because it relies on imaging modalities only that can differ in sensitivity and specificity^4,7^. There is an important need to develop sensitive tools to obtain an accurate snapshot of the tumor burden and identify true oligometastatic cancer patients who can benefit the most from SABR.

Alongside with circulating tumor cells and circulating tumor DNA, extracellular vesicles (EVs) have recently emerged as potential markers of tumor burden and predictors of response to therapy^30,35,36^. To detect EVs released from prostate cancer, we used two prostate-specific surface markers, namely PSMA and STEAP1, previously found enriched in prostate cancer cells and EVs^37,38^. PSMA has recently gained a lot of attention with the development of PSMA-directed radionuclide therapy and PET imaging^39,40^. Similarly, STEAP1 is currently under investigation as molecular imaging marker and therapeutic target^41,42^. Prior studies have reported the detection of PSMA and STEAP1 on surface of prostate cancer-derived EVs *in vitro* and in patient blood^38,43,44^. Interestingly, our study identifies STEAP1 as a robust marker of tumor-derived EVs in castration-refractory prostate cancer. At the time of diagnosis, we observed a strong correlation between levels of PSMA^+^-EVs and STEAP1^+^-EVs in CRPC patients. We also found that circulating STEAP1^+^-EVs outnumbered PSMA^+^-EVs in oCRPC and mCRPC patients. While antibody affinity can impact on EV quantification, it may also indicate differential expression of PSMA and STEAP1 in CRPC patients. Low PSMA expression has been previously reported in treatment-refractory patients^45–47^. In response to androgen-deprivation therapy, CRPC tumors can progress towards a neuroendocrine phenotype and lose PSMA expression^48–50^. Noteworthy, we detected PCEVs in all patients including those with undetectable PSA at baseline. Intratumoral and intertumoral heterogeneity of PSMA expression observed in advanced prostate cancer may have important consequences for patient selection using PSMA PET imaging and treatment with Lutetium-PSMA^45^. In a phase I study evaluating the safety profile of an antibody-drug conjugate targeting STEAP1, 99% of patients (133/134) showed positive STEAP1 expression and 73% with mid-high intensity^42^. Herein, we provide blood-based evidence that STEAP1 can be a promising alternative to PSMA for identification of aggressive prostate cancer including those characterized by neuroendocrine features and low serum PSA levels. In line with this, a recent study in lung cancer found that STEAP1 is overexpressed in poorly differentiated neuroendocrine lung cancer (small cell lung carcinoma) compared to carcinoid tumors^51^. Further studies are warranted to determine PSMA and STEAP1 differential expression in metastatic prostate cancer lesions identified by PET imaging.

To improve the identification of oligometastatic prostate cancer patients, we determined the association of baseline PCEV levels and tumor burden assessed by ^11^C-Choline PET imaging. Surprisingly, we did not find any correlation between PCEV levels and imaging features but baseline levels of PSMA^+^-EVs and STEAP1^+^-EVs were independent predictors of distant failure. Patients with high levels of PCEVs were more at risk of developing distant metastases early. This suggests that ^11^C-Choline PET imaging may underestimate disease burden leaving undetected metastases untreated. A PCEV-based blood test has the potential to help refine the identification of true oligometastatic prostate cancer who will benefit the most from SABR therapy. Given the emergence of PSMA PET imaging, comparative studies are needed to further evaluate the potential of PCEV levels as a marker of disease burden in oligometastatic patients diagnosed with PSMA and/or Choline PET imaging.

Longitudinal analysis post-SABR revealed that blood PCEV levels are rapidly increased following treatment with a peak at Day 7. While irradiation has been shown to stimulate EV biogenesis with *in vitro* cell cultures, our study is, to our knowledge, the first clinical demonstration of SABR-induced EV release^52–54^. Levels of PSMA^+^-EVs at Day 7 post-SABR were a strong predictor of oncological outcomes. In contrast to baseline, high levels of PSMA^+^-EVs were associated with lower risk of disease progression and better survival. This suggests that increase in blood PSMA^+^-EV levels may not only reflect SABR-associated cancer cell death but also SABR-mediated antitumor immunity. Preclinical and clinical studies from our group underscored that expansion of peripheral tumor-reactive CD8 T cells post-SABR is essential for local and systemic tumor control^7,11,13,28^. Herein, we found that elevation of PCEV concentrations was positively correlated with levels of tumor-reactive CD8 T cells. Interestingly, highest correlation was observed with CD8 T cells expressing CX3CR1 chemokine receptor. CX3CR1 is expressed by effector CD8 T cells and high levels of peripheral CX3CR1^+^ CD8 T cells have been linked to better response to immune checkpoint blockade^34,55^. While these findings point towards a crosstalk between tumors and CD8 T cells mediated by EVs, the nature of the association remains elusive. The peak of PCEV levels (Day 7) preceded the increased frequency of tumor-reactive CD8 T cells (Day 14) which suggests that PCEVs may play an active role in SABR-induced antitumor immunity. Similarly, melanoma patients who responded to pembrolizumab had levels of Ki67^+^ PD1^+^ CD8 T cells positively correlated with blood-derived exosomal PD-L1^30^. Tumor-derived EVs can carry immunomodulatory molecules such as PD-L1, CD73, miRNAs and cytosolic DNA that either induce or inhibit antitumor immunity^29,31, 56–58^. Prostate cancer is characterized by a tumor immunosuppressive microenvironment and EVs can contribute to tumor immune escape^59–61^. At baseline, we found an inverse correlation between PCEV levels and tumor-reactive CD8 T cells which may indicate an inherent systemic immunosuppressive microenvironment driven by PCEVs. Following SABR treatment, a subset of oCRPC patients have durable response with no signs of disease progression after 2 years. This suggests that, in these best responders, SABR can alter the molecular composition of PCEVs towards an immunostimulatory phenotype resulting in abscopal response^52,62^. Conversely, limited control of distant metastases can be attributed to the release of immunosuppressive EVs that impair with T-cell priming and tumor cell killing. We and others have shown that PD-L1 and B7-H3 immune checkpoint molecules can be upregulated on surface of EVs in response to radiotherapy^63,64^. Additional studies should focus on characterizing the molecular composition of PCEVs and determine the impact of SABR on expression of immunoregulatory molecules. Furthermore, it is critical to decipher the molecular and cellular mechanisms involved in EV-mediated antitumor immunity in response to SABR using immunocompetent prostate cancer mouse models. This will pave the way for the design of biomarker-driven combination therapies improving SABR efficacy and patient outcome.

We recognize several limitations to this study. Firstly, our study may be underpowered as the cohort is small with only 79 patients with blood available and not all patients provided blood for each timepoint. This, accompanied by its retrospective design, may introduce selection bias that confounds the study results. In this study, we used nanoscale flow cytometry as a means of establishing proof-of-concept of the clinical utility of measuring circulating tumor-derived EVs for patient stratification and prediction of response to SABR. While this technology is not primarily designed for clinical use, it has already been implemented in several clinical studies including randomized controlled trials^65–67^. Nanoscale flow cytometry offers numerous advantages for developing EV-based blood tests. It allows for high throughput multiparametric detection of EVs of ∼150 nm and greater. In addition, this technology has minimal requirements with respect to pre-analytical steps necessary to obtain a robust EV-based assay. This is particularly important consideration as most studies evaluating the clinical benefit of extracellular vesicles are limited by cost and time-consuming pre-analytical procedures affecting data reproducibility and implementation in a clinic setting. Our blood test can be performed within 1 hour from time of blood collection to data analysis using basic laboratory equipment, highlighting the potential for using such technology in the clinical setting with broad feasibility.

## Conclusions

This hypothesis-driven study shed light on a new EV-based blood test that may improve identification of patients with oligometastatic castration-refractory prostate cancer that will benefit from SABR treatment. Additionally, it provides novel biological insights in the crosstalk between tumor cells and adaptive immune cells in response to SABR. Future endeavors would involve validation in larger patient cohorts and comprehensive profiling of the molecular cargo of prostate cancer-derived extracellular vesicles to refine risk prediction and identify potential therapeutic vulnerabilities to improve response to SABR.

## Data Availability

All data produced in the present work are contained in the manuscript

## Acknowledgments

This research was supported by NCI R01 CA200551 (SSP, VJL, EDK, and HD), departmental grant (FL) and generous benefactors (FL).

## Author Contributions

SSP, HD, EDK, GBJ and VL conceived the clinical study and obtained approvals, funding, and recruitment to the OPeRATIC trial. EJT, VL, GBJ collected and reviewed Choline PET/CT imaging data. FL, YK, AA, HZ and JK performed laboratory studies. FL, JQ, IT, HA and FA conducted statistical modeling and data analyses. FL and SSP wrote the initial manuscript and all authors read and approved the final manuscript. FL and SSP designed and supervised the study.

---

**Figure S1.**
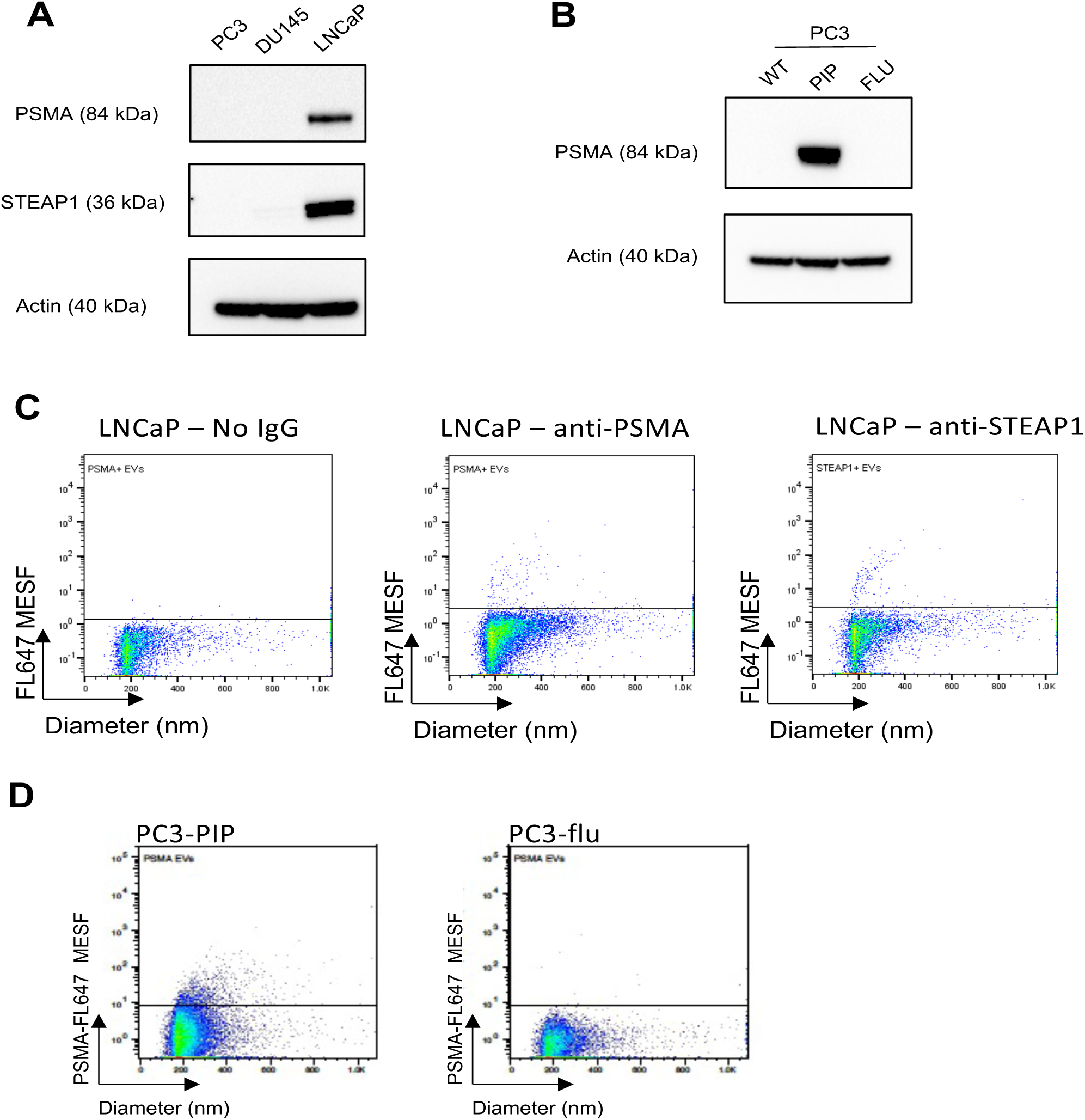
Nanoscale flow cytometric detection of PSMA^+^- and STEAP1^+^-EVs released from prostate cancer cells. A) Western-blot analysis of protein expression of PSMA and STEAP1 in prostate cancer cell lines B) Western-blot analysis of protein expression of PSMA in PC3 wild-type (WT), transfected with empty vector (FLU) and transfected with PSMA construct (PIP) C) Scatterplots showing nanoscale flow cytometric detection of PSMA- and STEAP1-positive EVs from LNCaP conditioned media D) Scatterplots showing nanoscale flow cytometric detection of PSMA- from PSMA-positive (PIP) and PSMA-negative PC3 cells

**Figure S2.**
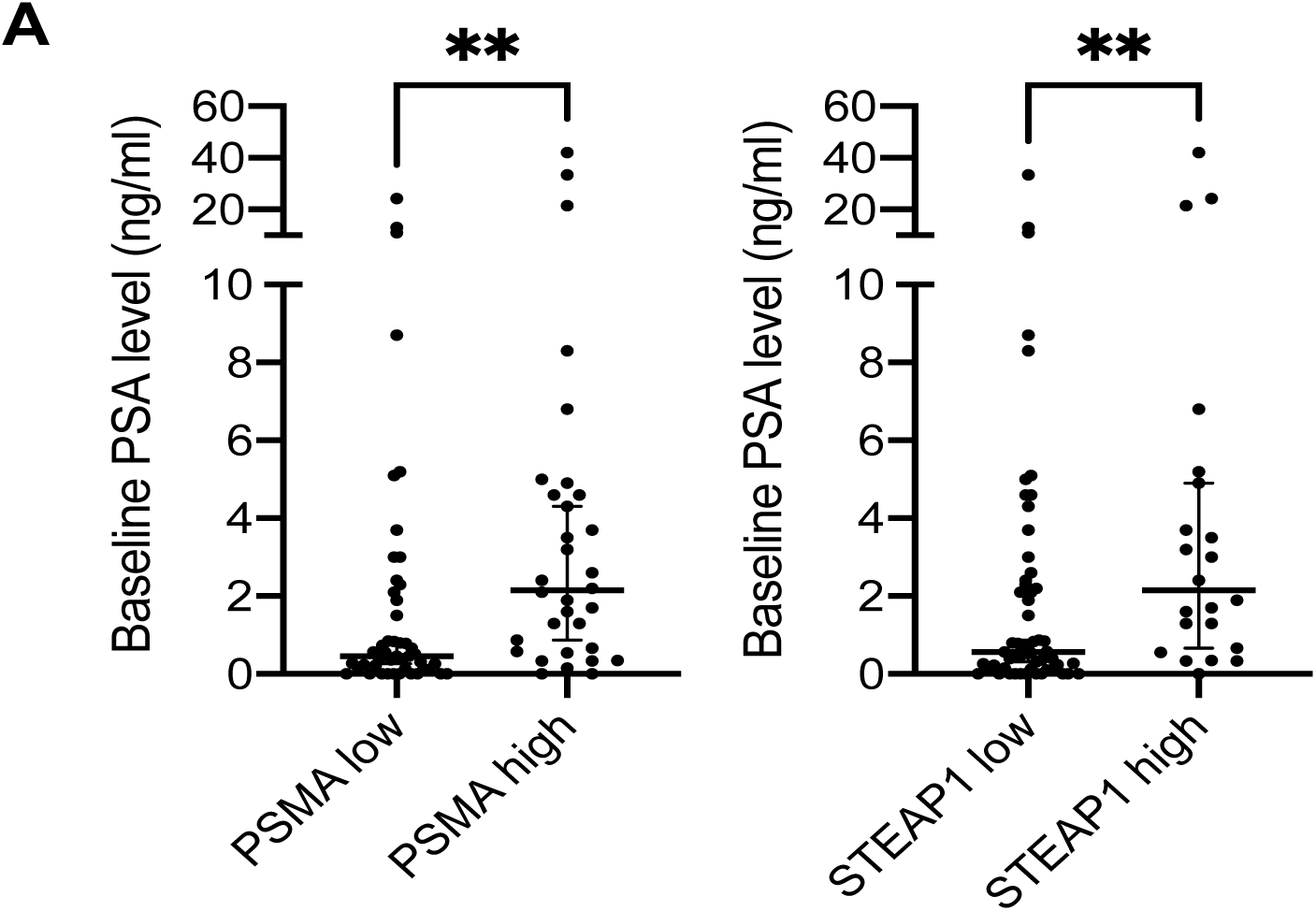
Association of serum PSA levels and baseline PCEV concentrations in omCRPC patients. A) Baseline PSA levels in omCRPC patients stratified by PSMA^+^- and STEAP1^+^-EV levels (n=79 patients total). Bars represent median and 95% Cl. Mann-Whitney U test, **p<0.01.

## Supplementary Materials

MIFlowCyt-EV List for standardized reporting of extracellular vesicles flow cytometry experiments

---

**Table S1.**
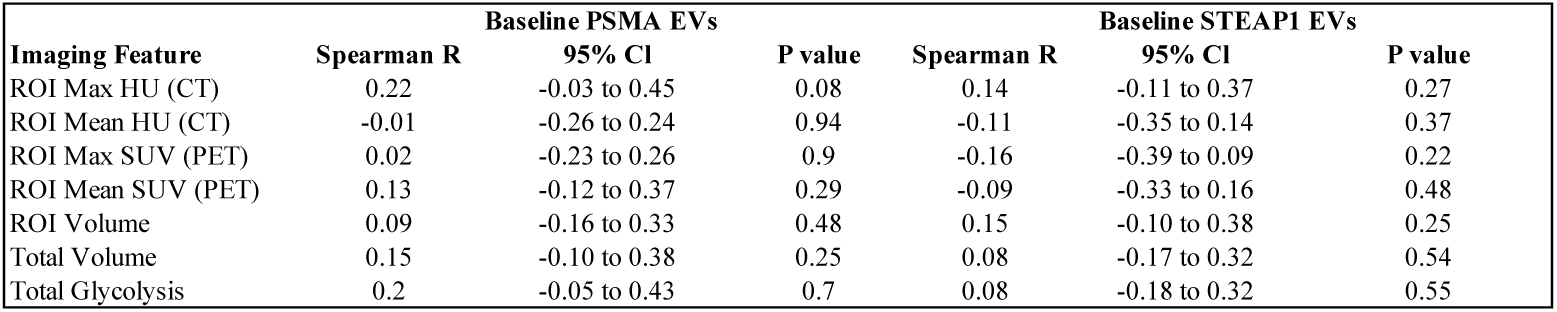
Correlation analysis between baseline PCEV levels and PET imaging features.

## MIFlowCyt-EV List

| Requirement | Please Include Requested Information |
| --- | --- |
| 1.1. Purpose | We intended to measure prostate cancer-derived extracellular vesicles in oCRPC patients treated with SABR to determine their clinical usefulness as prognostic and predictive biomarker. |
| 1.2. Keywords | Extracellular vesicles, nanoscale flow cytometry, prostate cancer, radiotherapy, |
| 1.3. Experiment variables | Pre-treatment and Day 1, 7, and 14 post-SABR |
| 1.4. Organization name and address | Mayo Clinic<br>Guggenheim 9-23<br>55901 Rochester, Minnesota<br>USA |
| 1.5. Primary contact name and email address | Fabrice Lucien-Matteoni, |
| 1.6. Date or time period of experiment | May 2020 – December 2020 |
| 1.7. Conclusions | This original study demonstrates that circulating PCEVs can serve as prognostic and predictive marker to SABR in order to identify “true” oCRPC patients. In addition, we provide novel insights in the global crosstalk mediated by PCEVs between tumors and immune cells which leads to systemic suppression of immunity against CRPC. This work lays the foundation for future studies that investigate the underpinnings of metastatic progression and providing new therapeutic targets (e.g PCEVs) to improve SABR efficacy and clinical outcomes in treatment resistant CRPC. |
| 1.8. Quality control measures | Fluorescence sensitivity was assessed using FITC and APC MESF beads (RCP-20-5, 0.4-0.6 μm, Spherotech). Side-scatter detection sensitivity and resolution was evaluated using Apogee bead mix (AB1493). Side-scatter intensities were converted in standardized units (nanometers) using Rosetta Calibration (Exometry, Amsterdam, The Netherlands). Staining of platelets with CD61-AF647 (VI-PL2 clone) was performed to determine presence of residual platelets in plasma. |
| 1.9 Other relevant experiment information | N/A |
| 2.1 Sample description |  |
| 2.1.1 Biological sample source description | Platelet-Poor Plasma from 79 oCRPC patients collected between 2016 and 2019 |
| 2.1.2 Biological sample source organism description | See table 1 for demographics and clinicopathological characteristics of patients. |
| 2.1.3 Environmental sample location | Samples were stored in cryovials at -80C |
| 2.2 Sample characteristics |  |
| 2.3. Sample treatment description | See ‘Blood and urine collection’ in the Materials and Methods section of the manuscript. |
| 2.4. Fluorescence reagent(s) description | Anti-PSMA and anti-STEAP1 monoclonal antibodies (see Materials and Methods section) |
| 3.1. Instrument manufacturer | Apogee, Northwood, UK |
| 3.2. Instrument model | A60-Micro Plus |
| 3.3. Instrument configuration and settings | Samples were analyzed at a flow rate of 1.5 μl/min on an A60-Micro Plus, equipped with a 405 nm laser (70 mW), 488 nm laser (70 mW) and 638 nm laser (70 mW). Samples were measured for 1 minute with triggering threshold on side scatter set at 23 a.u (cross-section scattering 9 nm<sup>2</sup>). AF488 fluorescence was collected in the 488-green channel (525/50 nm band pass filter). AF647 fluorescence was collected in the 638-red channel. PMT voltages were set to 320 V for SALS, 305 V for LALS, 500 V for 488-green and 450 V for 638-red. |
| 4.1. List-mode data files | Data were collected in fcs files |
| 4.2. Compensation description | No compensation needed |
| 4.3. Data transformation details | FlowJo (v10.6.2; FlowJo, Ashland, OR) and Rosetta Calibration (Exometry, Amsterdam, The Netherlands) were used to transform data acquired from Apogee Micro 60 Plus. |

